# Associations of Allopregnanolone and Related Steroid Hormones with Prenatal Psychosocial Distress in the Healthy Start Cohort

**DOI:** 10.64898/2026.07.22.26358684

**Authors:** Gabriella Mayne, K. Joseph Hurt, Sara Yeatman, Jost Klawitter, David P. Tracer, Uwe Christians, Dana Dabelea, Wei Perng

**Affiliations:** Department of Health & Behavioral Sciences, University of Colorado, Denver, CO, 80204 USA; Divisions of Maternal Fetal Medicine and Reproductive Sciences, Department of Obstetrics and Gynecology, University of Colorado Anschutz Medical Campus, Aurora, Colorado 80045, USA; iC42 Clinical Research & Development, Department of Anesthesiology, University of Colorado Anschutz Medical Campus, Aurora, CO 80045, USA; Lifecourse Epidemiology of Adiposity and Diabetes (LEAD) Center and the Department of Epidemiology, Colorado School of Public Health, University of Colorado Anschutz Medical Campus, Aurora, CO 80045, USA; Department of Pediatrics, School of Medicine, University of Colorado Anschutz Medical Campus, Aurora, Colorado 80045, USA

**Keywords:** Neuroendocrine, maternal health equity, prenatal mental health, precision medicine, social determinants of health

## Abstract

Psychosocial distress is associated with adverse perinatal outcomes, yet its biological correlates remain incompletely understood. We evaluated associations of prenatal psychosocial distress with allopregnanolone (ALLO) and related steroids, and assessed race/ethnicity as a potential effect modifier, using data from 237 participants from the Healthy Start Study. We selected participants based on high distress (Edinburgh Perinatal Depression Scale [EPDS] ≥13 or EPDS-3A ≥7; n = 57) or low distress (EPDS <4 and EPDS-3A <2; n = 180). We quantified ALLO, progesterone, pregnanolone, cortisol, and cortisone in maternal serum using HPLC-MS/MS at two timepoints in pregnancy for each participant, (median ∼17 and ∼27 weeks’ gestation; range: 10–34 weeks). We modeled the relationship between high vs. low prenatal distress with repeated measures of the steroid hormones using linear mixed-effects models. We found that ALLO was 20.6% lower (95% CI: –30.9%, –8.7%) in high-distress individuals after adjusting for maternal age, fetal sex, smoking and gestational age at blood draw, with no evidence of effect modification by race/ethnicity. However, this association attenuated after additional adjustment for sociodemographic characteristics. Several ALLO-related ratios were lower with high distress, but only ALLO-to-progesterone remained significant across models. These findings suggest that associations between psychosocial distress and circulating ALLO concentrations during pregnancy are influenced by social and structural factors, with the exception of the ALLO-to-progesterone ratio, which may capture aspects of neurosteroid metabolism more closely linked to prenatal psychosocial distress.

## INTRODUCTION

Prenatal psychosocial distress – reflected in part by depressive and anxiety symptoms occurring during pregnancy – affects roughly 20% of individuals,^1–4^ with higher rates among racial/ethnic minorities and individuals facing migration and economic deprivation.^5–7^ Prenatal distress is associated with pregnancy complications including substance use/abuse and suicide,^8^ low birthweight, preterm delivery, and impaired maternal-infant bonding.^9–13^ Pregnancy is a vulnerable period for maternal mental health, as many conditions labeled ‘postpartum’ actually emerge during gestation and may be more effectively addressed if identified early.^14^ Identifying biological correlates of prenatal psychosocial distress could clarify stress-responsive neuroendocrine pathways and may ultimately inform earlier detection and intervention to improve perinatal outcomes.^15^

Allopregnanolone (ALLO) is an inhibitory neuroactive steroid derived from progesterone, highly expressed during pregnancy, and a biologically plausible correlate of prenatal psychosocial distress.^16^ ALLO is stress-responsive, anti-inflammatory, and a positive allosteric modulator of gamma-aminobutyric acid (GABA).^17–20^ Depression and anxiety – key components of prenatal psychosocial distress – are increasingly understood as stress-related conditions characterized by inflammatory, neural-circuitry, and inhibitory–excitatory imbalances, with GABAergic signaling contributing to HPA-axis regulation.^21–23^ In animal models, chronic stress decreases circulating ALLO and produces anxiety and depression-like behaviors that are alleviated by exogenous ALLO administration.^17,24–29^ The efficacy of brexanolone and zuranolone highlights ALLO-linked GABAergic pathways as clinically relevant targets.^30–32^ However, the relationship between psychosocial distress and ALLO in pregnancy is less understood, even though ALLO levels rise to their highest endogenous concentrations^33–39^ and are thought to be crucial for maternal stress-regulatory and behavioral adaptations.^40,41^

Despite preclinical^17,24–29^ and pharmacological^42^ evidence suggesting that prenatal psychosocial distress may lower circulating ALLO levels, human studies in pregnancy are inconsistent. Some studies report lower ALLO among pregnant individuals with elevated psychosocial distress ^43,44^ while others find higher,^45–48^ non-linear,^39,49^ or null associations.^50–54^ These inconsistencies likely reflect the substantial variability across studies in design, the tools and thresholds used to define psychosocial distress, analytical assays, gestational timing, and cohort characteristics. They may also reflect differences in the nature and duration of stress exposure, as acute stress has been associated with transient increases in ALLO and chronic stress and stress-related disorders have been associated with reduced ALLO levels.^26^ Given ALLO’s relevance to stress physiology, clarifying the extent to which circulating ALLO is associated with prenatal psychosocial distress is an important step toward understanding the neuroendocrine pathways linking psychosocial distress and maternal mental health during pregnancy.

Building on emerging evidence linking neuroactive steroids to perinatal mental health, we evaluated whether maternal serum ALLO concentrations and related steroid measures including pregnanolone, progesterone, cortisol, cortisone, and their ratios with ALLO differed between pregnant individuals with high and low psychosocial distress. Additionally, because psychosocial distress is shaped by social and structural conditions,^55,56^ we tested whether associations between distress and the steroid measures differed by race and ethnicity, which we treat as social constructs reflecting differential exposure to these conditions. We hypothesized that pregnant individuals with high distress would have lower ALLO concentrations across gestation than those with low distress, and that these differences would be more pronounced among racial and ethnic minority groups. Across all objectives, we evaluated the direction, magnitude, and precision of associations across multiple biologically related steroid hormones involved in neurosteroid regulation and prenatal stress physiology. Examining patterns across related measures may provide a more comprehensive understanding of the physiological correlates of maternal psychosocial distress than consideration of any single measure alone.

## RESULTS

From the original Healthy Start pre-birth cohort of 1,410 individuals, 237 participants met inclusion criteria; 57 were classified as high-distress and 180 as low-distress (**Figure 1**). Compared with Healthy Start participants overall, the cohort in this study significantly differed in that it was ∼1 year older (29 vs. 28 years), less likely to be nulliparous (38% vs. 67%), less likely to use public assistance programs (31% vs. 39%), more highly educated (e.g., 27% vs. 20% with graduate degrees) and had higher annual household income (≥$70,000: 54% vs. 38%). Additionally, participants in this sample were more likely to identify as non-Hispanic White (60% vs. 43%) and reside in a rural area (3.5% vs. 1.1%; **Supplemental Table S1**).

**Figure 1.**
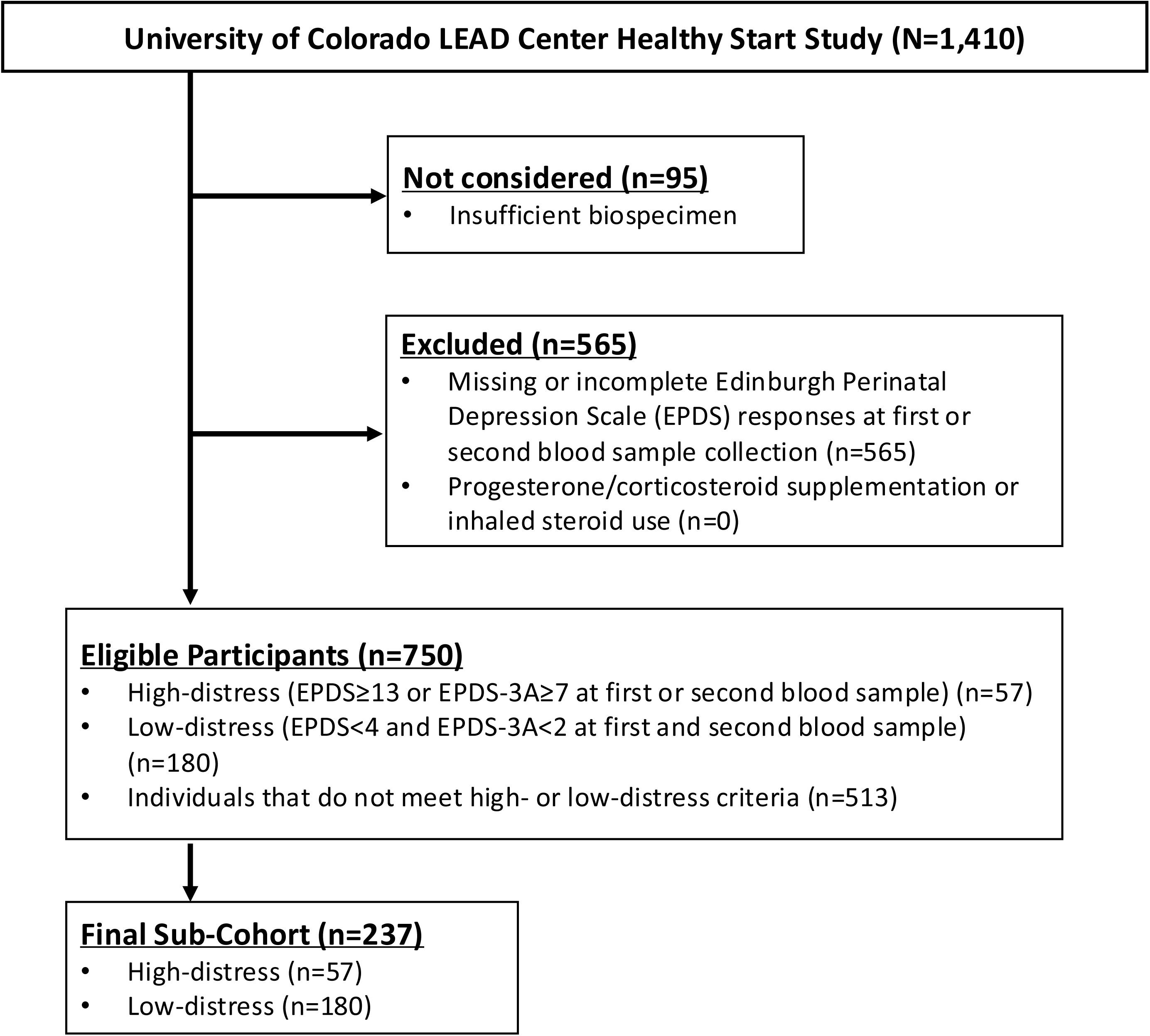
STROBE participant selection flow diagram for the present analysis, derived from participants in the Lifecourse Epidemiology of Adiposity and Diabetes (LEAD) Center Healthy Start Study.

As shown in **Table 1**, the analytic sample was 60% non-Hispanic White, 23% Hispanic, 11% non-Hispanic Black, and 6% non-Hispanic Other. Maternal age was 29 ± 5.9 years at the time of enrollment. In comparison to low-distress participants, those with high-distress were significantly younger (26 vs. 30 years), more likely to be single (32% vs. 11%), had lower educational attainment (<high school: 33% vs. 7%), more frequently used public assistance programs (58% vs. 22%), and were less likely to identify as non-Hispanic White (30% vs. 70%). High-distress participants were also less likely to be U.S.-born and more likely to smoke during pregnancy, be diagnosed with a current psychiatric disorder, and to use antidepressant or anti-anxiety medications (14% vs. 3%).

**Table 1.**
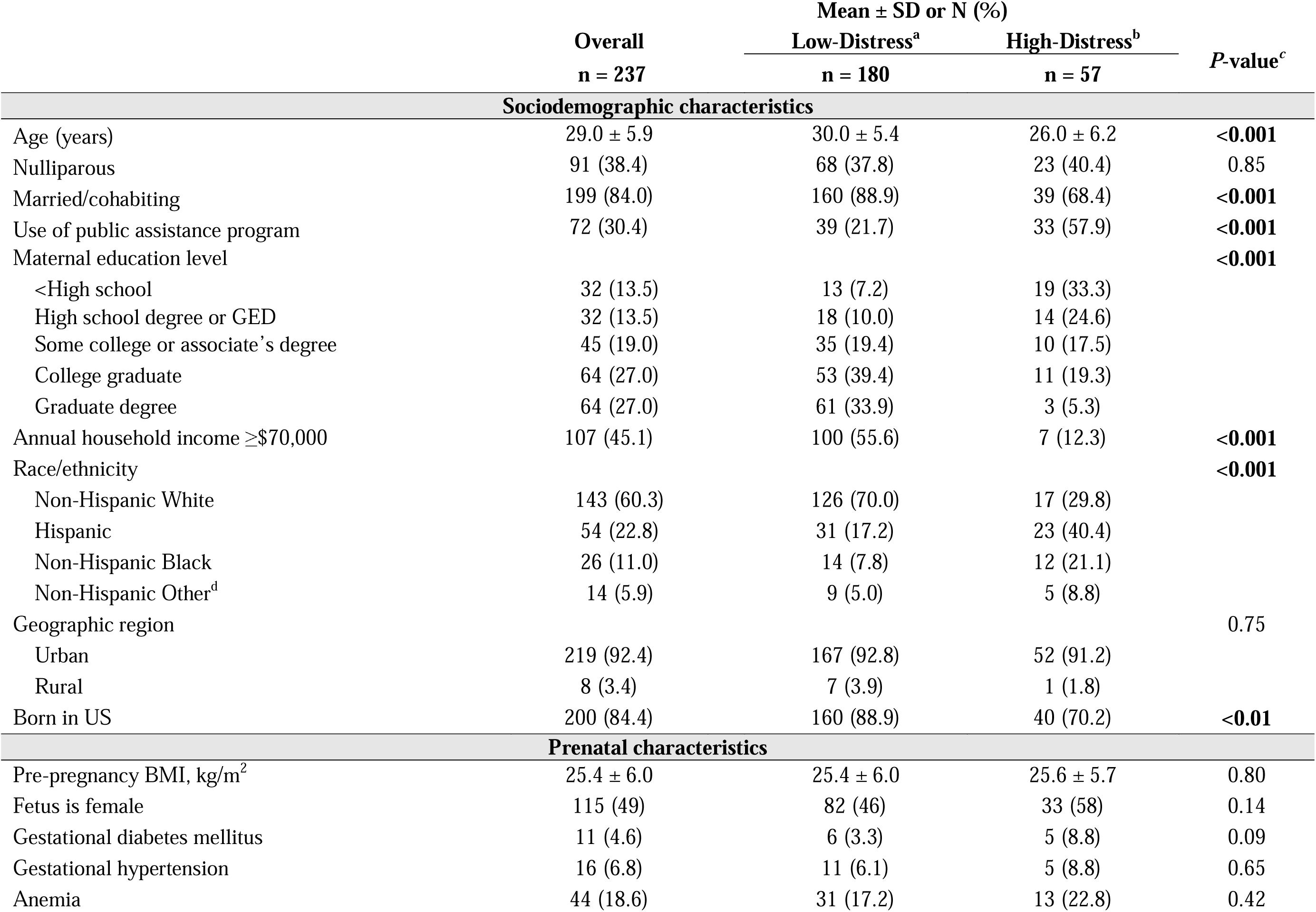
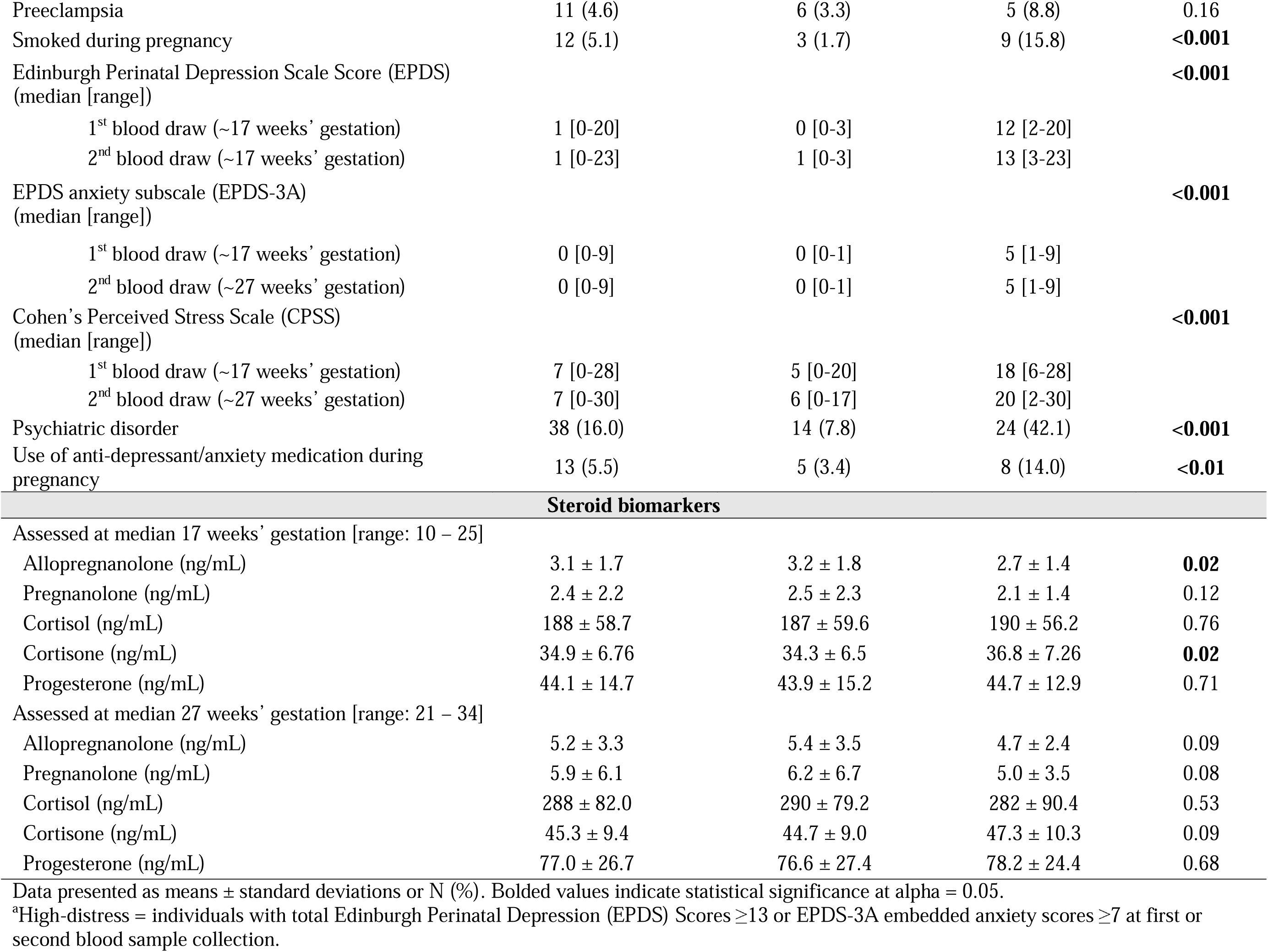
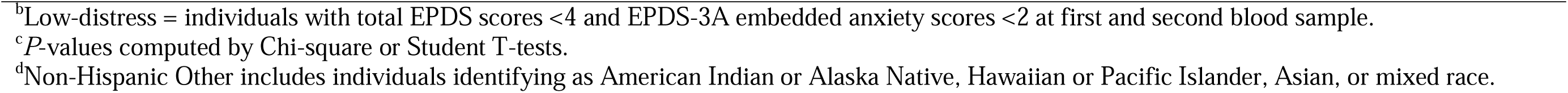
Bivariate associations of maternal sociodemographic and prenatal characteristics in the overall sample and stratified by high vs. low distress among 237 pregnant participants in the Healthy Start cohort.

**Table 2** shows the distribution of maternal serum ALLO levels at both pregnancy visits across sociodemographic and perinatal characteristics. ALLO levels were significantly lower among participants receiving public assistance and those with lower income. ALLO varied significantly by race/ethnicity at both visits and was lower among individuals born outside the U.S. ALLO was inversely associated with maternal pre-pregnancy BMI, positively associated with preeclampsia, and significantly higher among individuals with a male fetus.

**Table 2.**
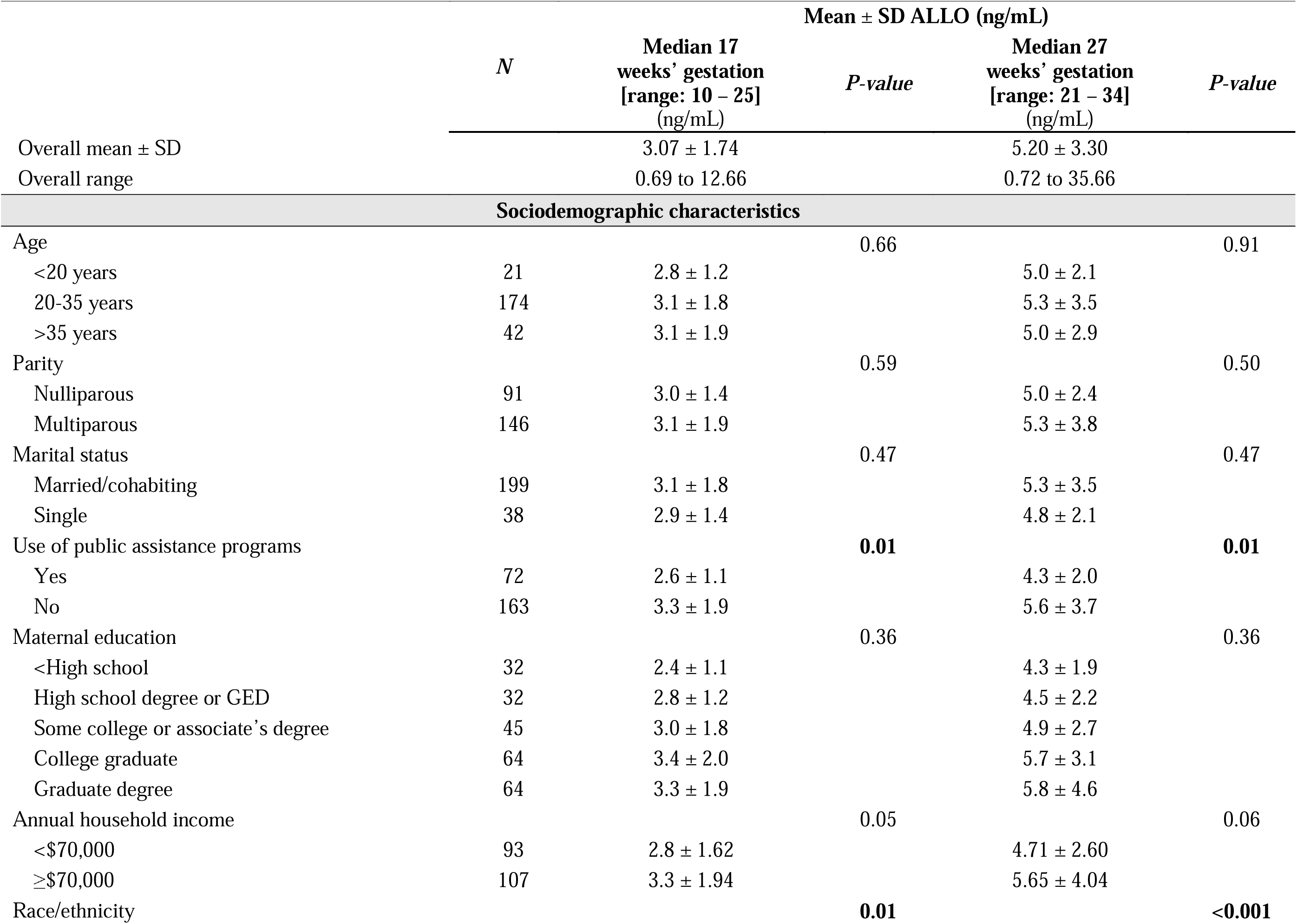
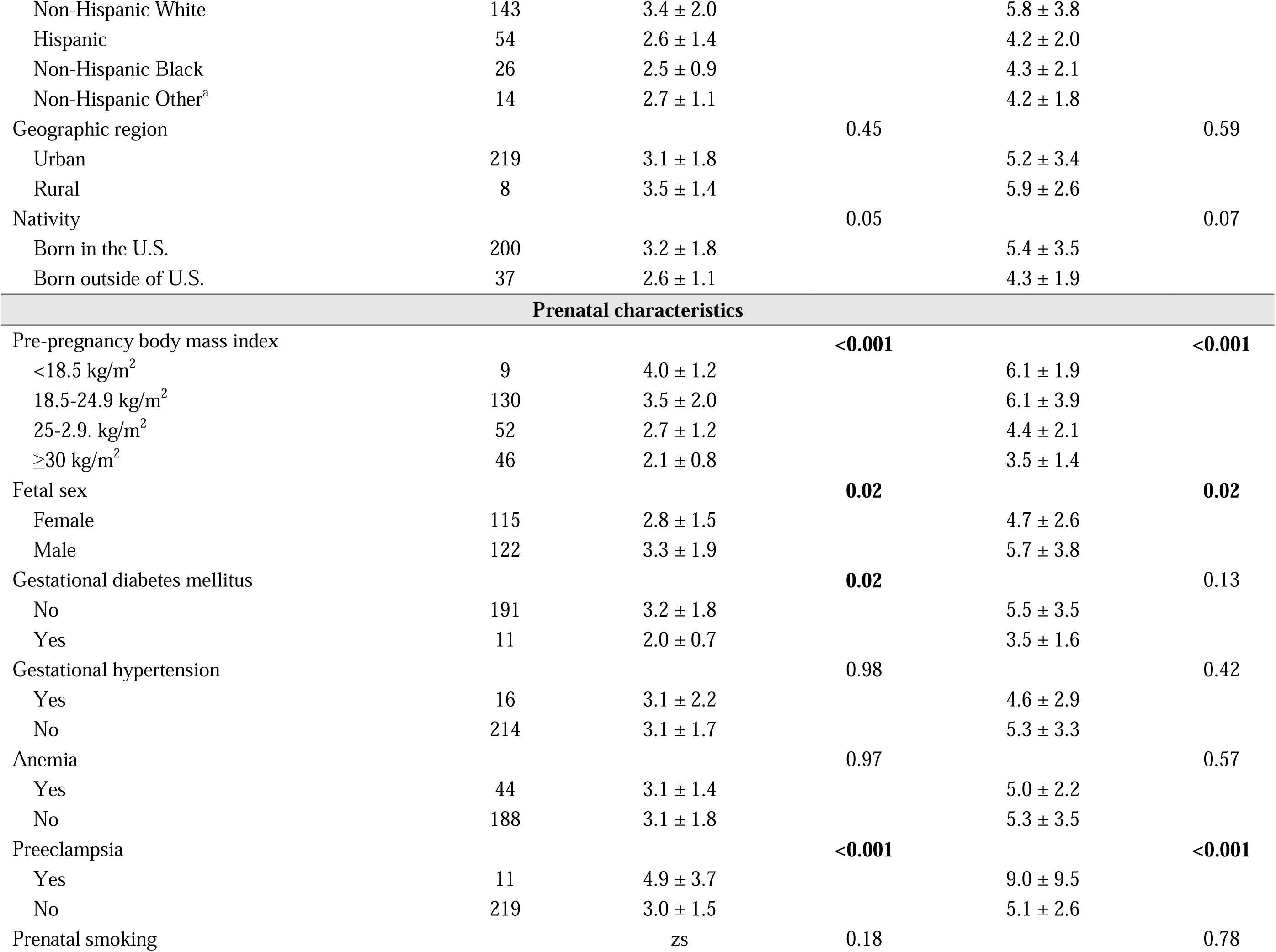
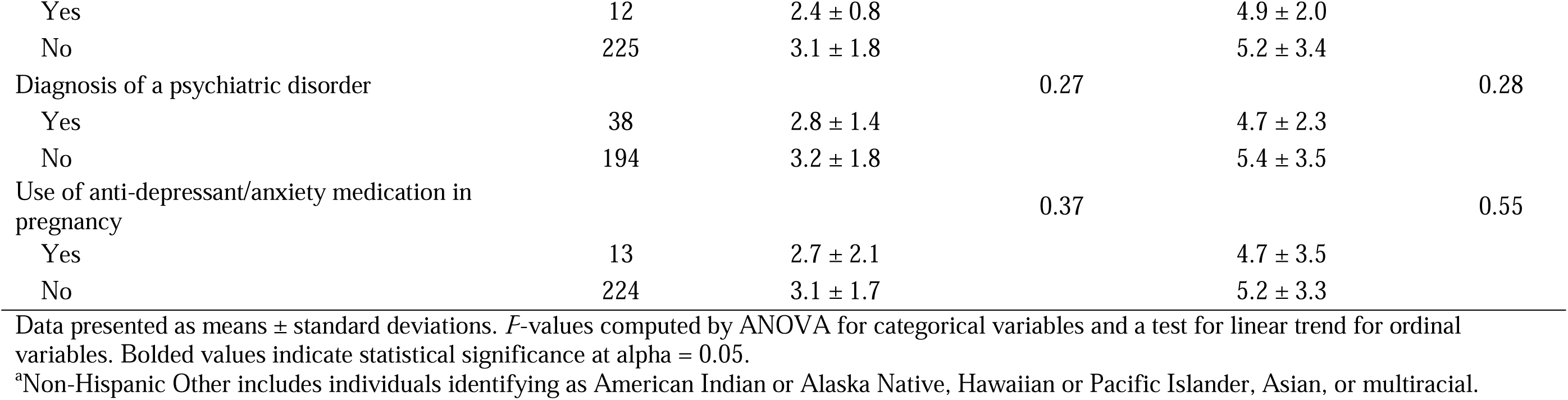
Bivariate associations of maternal sociodemographic and prenatal characteristics with allopregnanolone (ALLO) among 237 pregnant individuals in the Healthy Start cohort.

At both pregnancy timepoints, ALLO was moderately to strongly correlated with progesterone (rho = 0.59 at both visits) and pregnanolone (rho = 0.71 and 0.75), while cortisol and cortisone were also highly correlated with one another (rho = 0.66 at 17 weeks; rho = 0.61 at 27 weeks; **Supplemental Figure S1**). ALLO and glucocorticoids were moderately correlated across gestation, showing stronger alignment with cortisol at the first blood sample (∼17 weeks’ gestation) but a comparatively stronger association with cortisone at the second blood sample (∼27 weeks’ gestation) (**Supplemental Figure S1**). **Figure 2** displays the observed biomarker concentrations across gestation with trend lines illustrating raw data patterns by distress group. ALLO concentrations were lower in the high-distress group, aligning with the direction of effect estimates from the mixed-effects models.

**Figure 2.**
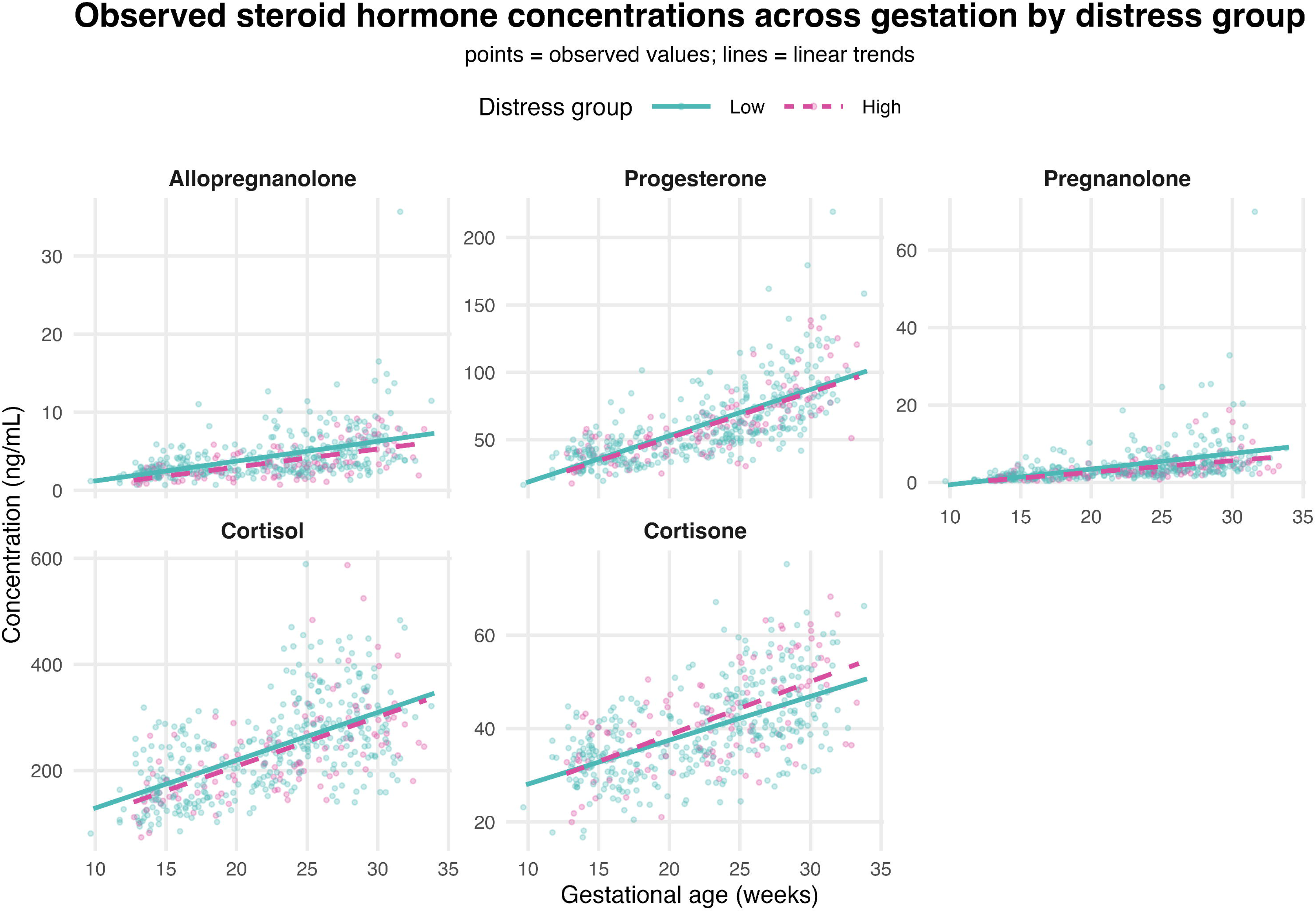
Observed steroid concentrations with linear fit lines showing low-distress (teal) and high-distress (pink) groups. One participant was an outlier for allopregnanolone (ALLO), pregnanolone, and progesterone at the second blood draw. This value appears biologically plausible. Sensitivity analyses removing the outlier (**Supplemental Table S3**) showed negligible changes to model estimates and therefore we retained the outlier in all analyses.

For the main analysis, in Model 1, which accounted for maternal age at enrollment, fetal sex, and prenatal smoking, individuals classified as having high psychosocial distress had 20.6% (95% CI: −30.9%, −8.7%) lower serum ALLO concentrations across gestation (observed range: 10 – 34 weeks) than those in the low-distress group (**Table 3**). Adjustment for sociodemographic factors (public assistance, nativity, and marital status) in Model 2 attenuated the estimate to approximately half the magnitude and the association was no longer statistically significant: −12.4% (95% CI: −24.4%, 1.4%). Adjustment for race/ethnicity in Model 3 further attenuated the estimate to −10.9%, 95% CI: −22.9%, 3.1%). We observed similar associations for pregnanolone, ALLO-to-cortisol, ALLO-to-cortisone, and cortisol-to-cortisone ratios, though these estimates were not statistically significant across models. In contrast, the ALLO-to-progesterone ratio remained lower in the high-distress group across multivariable models (**Table 3**). **Figure 3** provides a forest plot visualization of the mixed-effects model estimates, with full estimates in **Table 3**.

**Figure 3.**
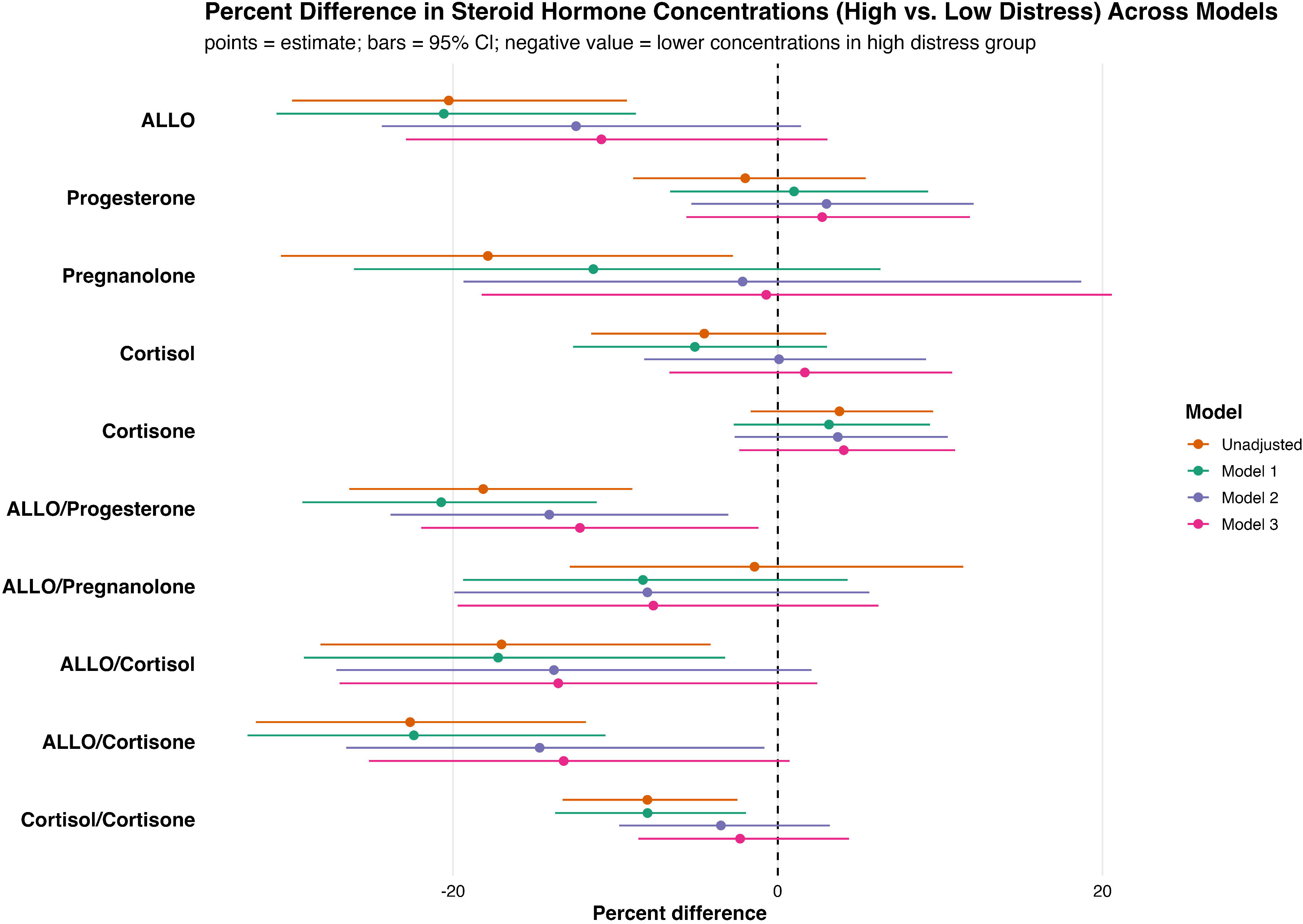
Forest plot of linear mixed effects model estimates (% difference) showing progressive attenuation of the associations across sequential models. Confidence intervals that do not cross zero indicate statistically significant differences between distress groups (*P* < 0.05). High-distress status is the fixed effect exposure of interest; fixed effects covariates in the unadjusted model included gestational age at blood draw and distress status only. Model 1 additionally adjusted for maternal age, fetal sex, and smoking status; Model 2 added public assistance use, marital status, and nativity. We observed no evidence of effect modification by race/ethnicity (*P* > 0.05) and therefore the fully adjusted Model 3 added race/ethnicity. We included random effects for individual intercepts for all steroids and steroid ratios; we included both individual intercepts and uncorrelated individual slopes for just ALLO, ALLO-to-progesterone, ALLO-to-cortisol, and ALLO-to-cortisone ratios based on improved model fit. Steroid concentrations were modeled on the log natural scale. % difference = (exp(estimate) – 1) *100. Full model estimates are provided in **Table 3**.

**Table 3.**
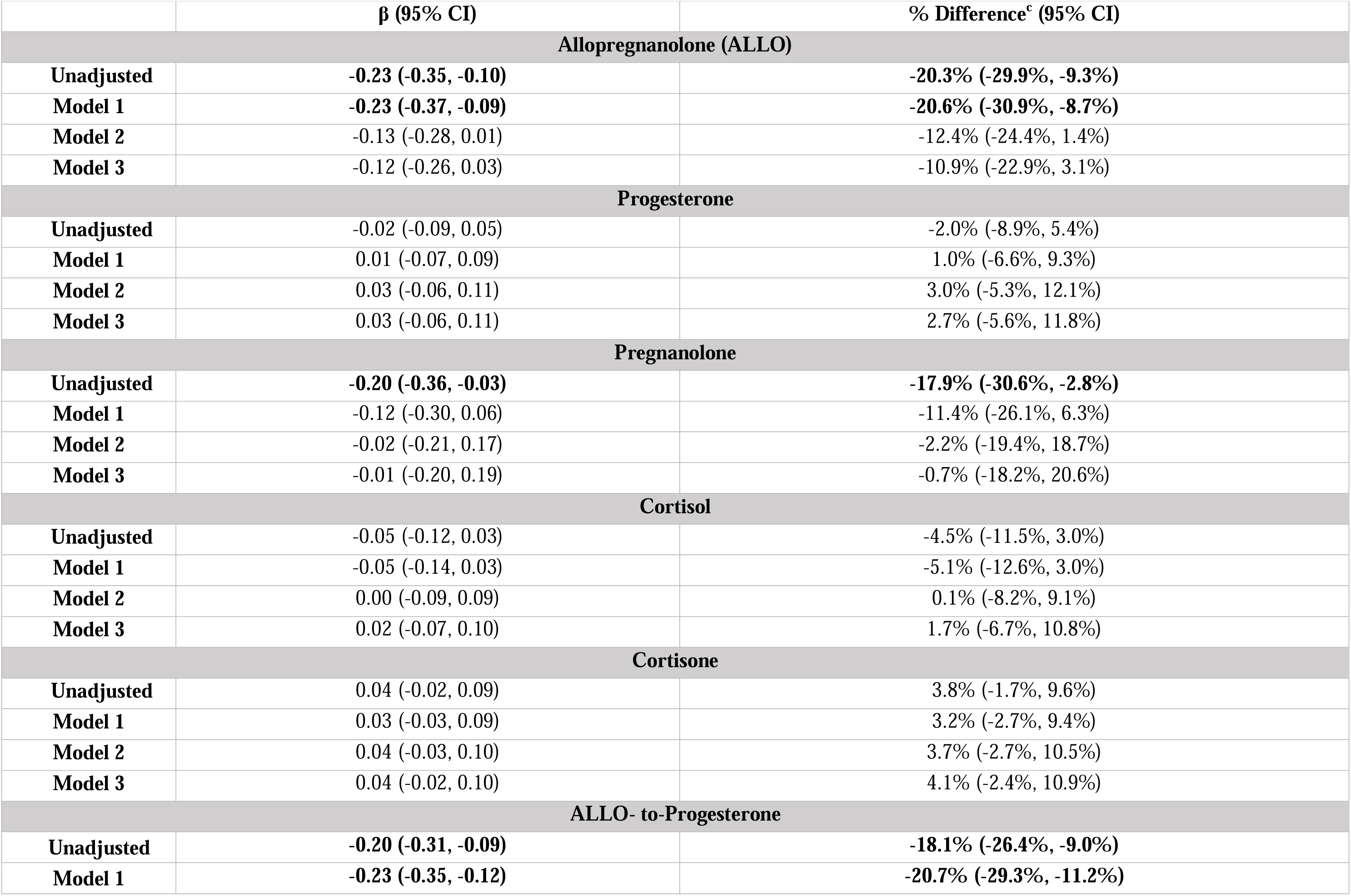
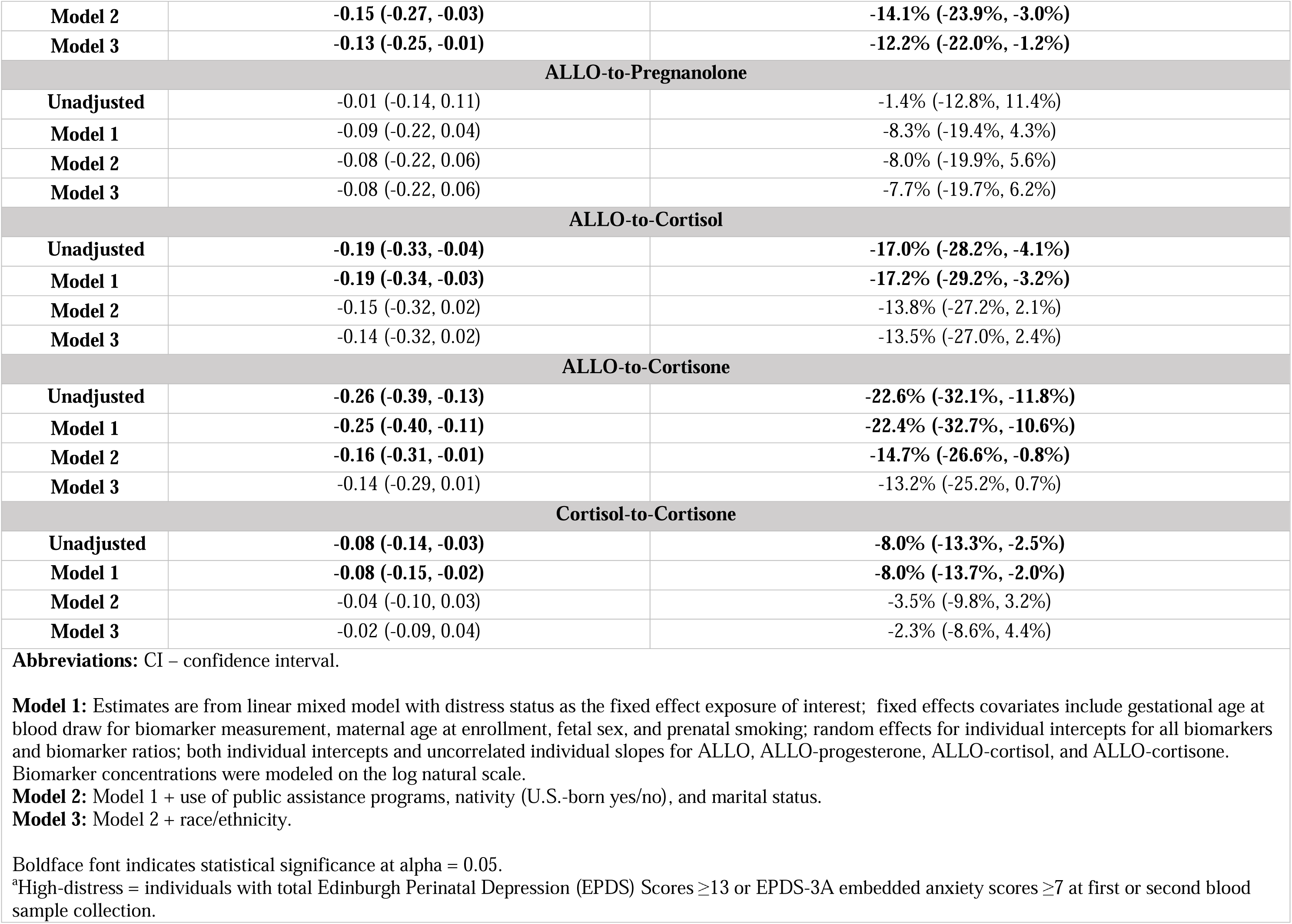
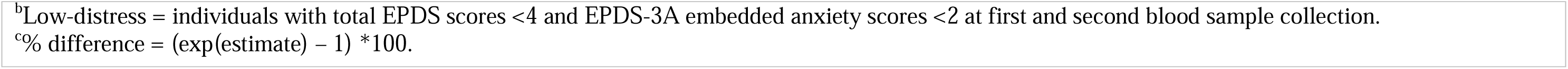
Average difference (β [95% CI] and % Difference [95% CI]) in natural log-(ln)-transformed allopregnanolone, related biomarkers, and allopregnanolone-biomarker ratios across 10 – 34 weeks’ gestation among 237 pregnant participants in the Healthy Start cohort with respect to high-distress^a^ (n = 57) vs. low-distress (n = 180)^b^

In sensitivity analyses, our findings were not altered by additional adjustment for maternal pre-pregnancy BMI, preeclampsia, or use of antidepressant/anti-anxiety medication (**Supplemental Table S2**), nor by the exclusion of a participant with outlying but biologically plausible values for ALLO, progesterone, and pregnanolone from the second blood draw (**Supplemental Table S3**). We also found no evidence that associations between psychosocial distress and maternal ALLO concentrations differed by race/ethnicity (Distress × Race/Ethnicity Type III interaction: χ² = 2.12, df = 3, *p* = 0.55); therefore, we did not pursue stratified models.

## DISCUSSION

In this study of 237 individuals from the Healthy Start pre-birth cohort, we found that maternal serum ALLO was ∼20% lower (∼0.5–1.0 ng/mL) for pregnant patients with high EPDS-defined psychosocial distress. However, this association attenuated after adjusting for sociodemographic characteristics and self-identified race/ethnicity, suggesting that both distress symptoms and neuroendocrine regulation may be shaped by shared upstream social and structural contexts. ALLO-to-progesterone ratios were also lower among individuals with high distress even after multivariable adjustment. Together, these findings suggest that neurosteroid metabolism may reflect the physiological manifestation of prenatal psychosocial distress.

### Prenatal psychosocial distress and ALLO

Our findings contribute to a growing but inconsistent body of human research on psychosocial distress and ALLO during pregnancy. Some studies report lower ALLO among distressed individuals, including Hellgren et al., who observed reduced term ALLO measured by celite chromatography/RIA among participants with high Montgomery–Åsberg scores,^43^ though no associations emerged earlier in pregnancy or when distress was defined using EPDS.^50^ Crowley et al. similarly found that greater negative affect and sleep disturbance corresponded to lower second-trimester ALLO+pregnanolone (immunoassay) in a small U.S. cohort.^44^ In contrast, other studies report higher ALLO among distressed groups.^45–48^ Wenzel et al. noted elevated early-pregnancy ALLO (LC–MS/MS) among individuals with first-onset depression but lower ALLO in those with recurrent depression in a predominantly Black and Hispanic cohort.^46^ Deligiannidis et al. reported higher plasma ALLO (LC–MS/MS) in a small, predominantly non-Hispanic White cohort (n=75) using narrower EPDS thresholds (EPDS ≤ 5 vs. EPDS ≥ 10).^48^ Still other studies report null associations,^50,51,53,54,57^ including a recent meta-analysis; however, its sensitivity analyses showed bidirectional patterns depending on antidepressant use and study quality, highlighting substantial heterogeneity across samples and methods.^52^ Emerging evidence also suggests that distress-ALLO associations may be non-linear,^39,49^ though the ‘U-shaped’ pattern observed in one study was from serial late-pregnancy salivary ALLO measurements using ELISA.^39^ Within this mixed literature, our study provides evidence that circulating ALLO concentrations are lower across 10-34 weeks’ gestation among individuals with EPDS-defined high psychosocial distress, but this association was no longer evident after accounting for social and structural factors. Importantly, we view this attenuation as informative, as it suggests that the relationship between psychosocial distress and neuroendocrine regulation during pregnancy is embedded within a broader ecosocial framework.^58^ Both ALLO regulation and psychosocial distress may reflect shared upstream social and physiological processes. From this perspective, circulating ALLO concentrations may reflect not just psychosocial distress itself, but also the social and structural conditions through which distress becomes embodied.^59^

Inconsistent findings regarding prenatal psychosocial distress and circulating ALLO in the literature likely reflect several factors, including differences in neurosteroid regulation, acute vs. chronic stress, assay rigor, and cohort diversity. Experimental and clinical studies demonstrate that GABA_A_ receptor plasticity, subunit composition, and sensitivity are central to mood physiology.^21,60^ From this perspective, both elevated and insufficient neurosteroid signaling – reflecting hyper- or hypo–stress reactivity – may indicate distress,^61^ or could also reflect tolerance-related changes in GABA_A_ receptor sensitivity.^62^ A large meta-analysis further showed that assay type influences ALLO estimates;^63^ accordingly, we used HPLC-MS/MS to avoid the cross-reactivity and limited specificity associated with ELISA and radioimmunoassay methods.^64^ Population differences may also contribute to heterogeneous findings. Cohorts differ widely in lifetime exposures to social and structural adversity, underscoring the need for socio-demographically diverse cohorts when interpreting ALLO as a potential biological correlate of prenatal psychosocial distress. In our study, ALLO varied by sociodemographic characteristics, and adjusting for these factors markedly attenuated the association between psychosocial distress and ALLO – highlighting the role of upstream social determinants in shaping both perceived distress and neuroendocrine profiles. Consistent with this, a separate study found lower baseline ALLO and reduced stress reactivity among non-pregnant African American women compared with White women,^65^ suggesting that differential exposure to chronic social stressors and structural inequities may influence ALLO regulation even outside of pregnancy.^66^ Antidepressant use may also contribute to heterogeneity across studies, as selective serotonin reuptake inhibitors (SSRIs) can influence neurosteroid signaling independent of depressive symptom severity.^67,68^ In our cohort, however, additional adjustment for antidepressant and anti-anxiety medication use did not materially alter the results.

### Secondary analyses of steroid hormones and ratios

In secondary analyses, we observed no associations between distress with cortisol and cortisone individually. However, prenatal distress was associated with a lower cortisol-to-cortisone ratio in the unadjusted model and Model 1, suggesting that glucocorticoid metabolism may be altered even when absolute concentrations are not. This finding is notable because prior studies, including our own in the Healthy Start cohort^69^ – have reported altered cortisol-to-cortisone ratios in preterm birth.^70^ The inverse association was similar for ALLO-to-cortisol and ALLO-to-cortisone ratios, though these estimates became attenuated and the 95% CI included the null after adjustment for the sociodemographic characteristics and race/ethnicity, suggesting that social and structural factors contribute to both psychosocial distress and glucocorticoid regulation during pregnancy. While these ratios were not independently associated with distress after accounting for these factors, the pattern of attenuation may help identify upstream targets to improve maternal well-being.

On the other hand, while the association of high prenatal distress with lower ALLO-to-progesterone ratios was similarly attenuated with multivariable adjustment, the estimate remained statistically significant, suggesting that prenatal distress may be associated with reduced enzymatic conversion of progesterone to ALLO. This finding aligns with stress-related downregulation of 5α-reductase and 3α-hydroxysteroid dehydrogenase^71^ – key enzymes in ALLO biosynthesis. These findings also appear to align with findings from a study assessing the ALLO-to-progesterone ratio in mid and late pregnancy in individuals with anxiety^72^ but contradict a separate finding of elevated ratios in depressed pregnant patients.^46^ The persistence of the association across models suggests the relationship between ALLO and one of its key precursors may provide information beyond absolute hormone concentrations alone.

Recent FDA approval of a digital therapeutic application that improves EPDS scores highlights the continued relevance of validated self-report tools in perinatal mental health.^73^ In our study, distress-related differences in Cohen’s Perceived Stress Scale (CPSS) scores – despite not being used for group classification – support that EPDS captures broader dimensions of psychosocial distress.^74^ These findings point to several exciting research avenues, including clarifying the causal direction between psychosocial distress and neurosteroid regulation – which may be bi-directional^49^ – and testing whether interventions that buffer or reduce distress alter ALLO concentrations and ALLO-to-progesterone ratios. Future work could also examine whether integrating neurosteroid measures with validated self-report instruments provides additional insight into the physiological correlates of prenatal psychosocial distress. The persistence of lower ALLO-to-progesterone ratios across fully adjusted models suggests that markers of neurosteroid metabolism may provide complementary information beyond absolute ALLO concentrations alone. Finally, this line of research may help identify therapeutic targets, including GABAergic signaling – an area under active investigation with potential future relevance in pregnancy.^22,75^

### Strengths and limitations

This study has several strengths. Use of a validated, highly specific HPLC-MS/MS assay enabled precise quantification of ALLO, its precursors, and glucocorticoids. Our clear group separation enhanced biological interpretability and reduced misclassification. Longitudinal modeling leveraged repeated sampling and allowed inference on the association between psychosocial distress and ALLO and other steroid hormone levels across a broad portion of pregnancy. Finally, our rich covariate data enabled us to model the impact of social and structural covariates that are increasingly recognized as drivers of pregnancy and maternal/infant health.

This study also has several limitations. First, to ensure adequate statistical power, we defined high-distress as EPDS ≥13 or EPDS-3A ≥7 at either pregnancy visit. As a result, we could not evaluate associations of chronicity or changes in distress symptoms over pregnancy. This also limits inference about the temporal relationship between psychosocial distress and ALLO. Second, small sample sizes limited disaggregation of racial and ethnic groups – particularly for American Indian and Alaska Native participants who face higher rates of perinatal distress^76^ and maternal mortality,^77^ yet remain systematically underrepresented in maternal health research.^78–80^ Third, we did not evaluate isopregnanolone (3β,5α-THP), a stereoisomer of ALLO, during assay method development or validation. Therefore, we cannot exclude the possibility that this neuroactive steroid contributed to the measured ALLO signal or affected quantification. However, all samples were analyzed using the same assay and analytical conditions, making it less likely that any such interference would systematically differ by distress status and explain the observed group differences. Fourth, because our analytic sample was enriched for individuals at the extremes of EPDS scores, the generalizability of these findings to the broader population may be limited. Finally, as with all observational studies, we cannot rule out residual confounding, and unmeasured factors may account for some of the observed differences.

## Conclusion

In sum, we found lower circulating ALLO concentrations among individuals with high prenatal psychosocial distress, and these associations were largely explained by social and structural factors. We also observed that prenatal distress associated with lower values for glucocorticoid ratios and ALLO-to-glucocorticoid ratios, but with only the ALLO-to-progesterone ratio consistently associated with distress across multivariable models. The persistence of lower ALLO-to-progesterone ratios suggests that markers of neurosteroid metabolism may provide complementary information beyond absolute ALLO concentrations alone. Taken together, our findings suggest that neurosteroid regulation during pregnancy reflects the interplay of social, structural, and physiological processes that shape maternal adaptation.

## Data Availability

De-identified participant data and analytic code supporting the findings of this study are available from the corresponding author upon reasonable request and with approval from the Healthy Start Study and the Colorado Multiple Institutional Review Board.

## Resource Availability

### Lead Contact

Further information and requests for resources should be directed to and will be fulfilled by the lead contact, Gabriella Mayne.

### Materials Availability

This study did not generate new unique reagents.

## Acknowledgments

The Healthy Start Study is supported by the National Institutes of Health (R01 DK076648). This analysis received no external funding. Institutional support from iC42 Clinical Research and Development and the Department of Health and Behavioral Sciences (University of Colorado Anschutz/Denver) contributed to assay implementation, data analysis, and manuscript preparation. Funders had no role in study design, data interpretation, or the decision to submit the manuscript. We thank the LEAD Center biorepository staff for biospecimen preparation and the iC42 research specialists for their assistance with the bioassays.

## Author Contributions (CRediT taxonomy)

G.M.: Conceptualization, Data Curation, Formal Analysis, Investigation, Methodology, Visualization, Writing – Original Draft. W.P.: Conceptualization, Methodology, Formal Analysis, Supervision, Validation, Writing – Review & Editing. D.T.: Conceptualization, Methodology, Writing – Review & Editing. S.Y.: Conceptualization, Methodology, Writing – Review & Editing. K.J.H.: Conceptualization, Methodology, Supervision, Writing – Review & Editing. J.K.: Investigation, Methodology, Software, Validation, Writing – Review & Editing. D.D.: Resources, Supervision, Writing – Review & Editing. U.C.: Conceptualization, Methodology, Resources, Supervision, Validation, Writing – Review & Editing. G.M. and W.P. accessed and verified the underlying data. All authors critically revised the manuscript and approved the final version.

Declaration of Generative AI and AI-assisted Technologies in the Writing Process During the preparation of this work, the authors used ChatGPT (OpenAI) in order to improve the readability and language of the manuscript. After using this tool, the authors reviewed and edited the content as needed and take full responsibility for the content of the published article.

## STAR METHODS

### KEY RESOURCES TABLE

| RESOURCE | SOURCE | IDENTIFIER |
| --- | --- | --- |
| Healthy Start Study cohort | Healthy Start Study | COMIRB #09-0563 |
| Edinburgh Perinatal Depression Scale (EPDS) | Cox et al., 1987 | PMID: 3651732 |
| HPLC-MS/MS steroid assay | Mayne et al., 2021 | PMID: 34279681 |
| Allopregnanolone standard | Toronto Research Center | Catalog #A547100 |
| Progesterone standard | Cayman Chemicals | Catalog #15876 |
| Pregnanolone standard | Sigma Aldrich | Catalog #P8129 |
| Cortisol standard | Cayman Chemicals | Catalog #20739 |
| Cortisone standard | Toronto Research Center | Catalog #C696500 |
| Allopregnanolone-d5 internal standard | Sigma Aldrich | Catalog #809853 |
| Progesterone-d9 internal standard | Sigma Aldrich | Catalog #P-070 |
| Pregnanolone-d5 internal standard | Toronto Research Center | Catalog#P712203 |
| Cortisone-d8 internal standard | Sigma Aldrich | Catalog #900170 |
| Cortisol-d4 internal standard | Sigma Aldrich | Catalog #C-113 |
| R software (v4.5.1) | R Foundation for Statistical Computing | https://www.r-project.org |
| lme4 package | CRAN Repository | https://CRAN.R-project.org/package=lme4 |
| lmerTest package | CRAN Repository | https://CRAN.R-project.org/package=lmerTest |

### Experimental Model and Study Participant Details

#### Study population and psychosocial distress assessment

All participants provided written informed consent. The Colorado Multiple Institutional Review Board approved the Healthy Start Study and this secondary analysis (COMIRB #09-0563 and COMIRB #24-0224).

We performed a retrospective cohort study using biobanked specimens from the Healthy Start Study, a prospective pre-birth cohort of 1,410 pregnant individuals recruited at the University of Colorado Anschutz Medical Campus (Aurora, CO, USA) between December 2009 and May 2014.^81^ Eligible participants were ≥16 years old, had a singleton pregnancy, were ≤ 23 weeks’ gestation at enrollment, and reported no history of cancer, psychiatric disease, steroid-dependent asthma, preexisting diabetes mellitus, prior preterm birth, prior low birthweight delivery, or fetal demise. Twice during pregnancy (median 17-and 27-weeks’ gestation), participants provided morning fasting venous blood samples and completed mood and behavior questionnaires (Edinburgh Perinatal Depression Scale (EPDS) and Cohen’s Perceived Stress Scale (CPSS). Blood samples were collected by venipuncture and processed according to Healthy Start Study biorepository protocols. Serum was separated within one hour of collection, aliquoted into polypropylene cryovials in 0.5 mL aliquots and stored at −80 °C for future research use.^82^ Serum specimens used in the present study were collected between 2009 and 2014 and remained in long-term frozen storage (10 – 15 years) until neurosteroid quantification. Prior to analysis, samples underwent no more than three freeze-thaw cycles associated with specimen retrieval and previous research use.

For this study, we excluded participants who used progesterone, corticosteroids, or steroid inhalers during pregnancy, those with insufficient stored serum, and those for whom we did not have complete EPDS data. We selected participants based on distress status (high vs. low) using EPDS scores and the three-item anxiety subscale (EPDS-3A).^83,84^ We classified participants as high-distress if they had EPDS ≥ 13 and/or EPDS-3A ≥ 7 at either visit, reflecting clinically meaningful depressive or anxiety symptoms.^85–89^ We classified participants as low-distress if they had EPDS < 4 and EPDS-3A < 2 at both visits, indicating minimal or no symptoms.^85,86^ We focused on the EPDS as it is a widely used, clinically validated, and cross-culturally translated screening tool for depression and anxiety in perinatal care.^83,85,88,90–93^ We intentionally employed an “extreme phenotyping” approach to sharpen biological contrast and improve our ability to detect differences in steroid profiles.^94,95^ We calculated a sample size of 170 individuals using previously reported differences in third-trimester ALLO levels between depressed and non-depressed pregnant individuals (10.8 ± 5.7 vs. 17.4 ± 6.0 ng/mL)^50^ assuming 95% power and an error rate of 0.01. Because our ALLO measurements were taken earlier in pregnancy and quantified using a more specific assay (HPLC-MS/MS) – both factors expected to yield lower absolute concentrations – we retained all individuals who met criteria for high and low distress to avoid selection bias. This approach yielded a final analytic sample of 237 participants, 57 of whom were classified as high-distress and 180 as low-distress.

### Method Details

#### Neurosteroid and steroid hormone quantification

We quantified allopregnanolone (ALLO; 3α-hydroxy-5α-pregnan-20-one), progesterone, pregnanolone (3α-hydroxy-5β-pregnan-20-one), cortisol, and cortisone in maternal serum using a previously published validated high-performance liquid chromatography–tandem mass spectrometry (HPLC–MS/MS) assay developed to measure ALLO and related steroid hormones during pregnancy.^69,96^ Additional details on sample preparation and assay performance are provided in **Supplemental Methods.**

All chemicals, reagents, buffers, and columns were identical to the original assay with a minor modification to the HPLC gradients following transfer to the analytical platform used in the present study. Briefly, extracts were loaded onto the guard column using 85% aqueous buffer and 15% organic buffer. During the 1-minute loading period, the flow rate was increased from 800 μL/min to 1500 μL/min. After one minute, the switching valve was activated, and the retained analytes were backflushed onto the analytical column using 35% aqueous buffer and 65% organic buffer at a flow rate of 800 μL/min. This composition was kept constant for 4 minutes. Thereafter, the organic buffer was increased to 70%, after 6.5 minutes it increased to 77%, after 8 minutes to 83%, after 8.5 minutes to 88%, and after 8.8 minutes to 100%. This was followed by re-equilibration to the starting conditions at minute 8.9 and the assay was complete at 10 minutes. Before analyzing study samples, we conducted a three-day abbreviated validation on this platform pursuant to clinical and industry guidelines.^97^ The purpose of this validation was to confirm the assay performance following transfer of the previously validated method. The assay was successfully transferred to the new platform and met acceptance criteria. Intra- and inter-day accuracy and precision were within predefined acceptance limits (**Supplemental Tables S4**). The working ranges of the assay on the present platform were 0.39–100 ng/mL for ALLO and pregnanolone, 1.56–400 ng/mL for progesterone and cortisone, and 3.91–1000 ng/mL for cortisol. Representative extracted ion chromatograms and method details, including steroid standards, internal standards, and mass spectrometer parameters, are provided in the Key Resources Table as well as in **Supplemental Tables S4-S5, Supplemental Figures S2-S3,** and **Supplemental Methods**.

### Covariates

We obtained maternal characteristics from prenatal questionnaires and medical records, including age, marital status, educational attainment, annual household income, public assistance use (SNAP or WIC), pre-pregnancy BMI (kg/m²), prenatal cigarette smoking, and fetal sex. Maternal race and Hispanic ethnicity were self-reported using standard categories (American Indian or Alaska Native, Asian, Black, Native Hawaiian or Pacific Islander, White, multiracial, other; Hispanic ethnicity yes/no). For analysis, we grouped participants into mutually exclusive, census-derived categories: non-Hispanic White, non-Hispanic Black, non-Hispanic Asian, non-Hispanic Other (including American Indian or Alaska Native, Native Hawaiian or Pacific Islander, multiracial individuals, and those not otherwise specified), and Hispanic (any race).^98^ We also obtained pregnancy and birth characteristics, including parity (nulliparous/multiparous), gestational diabetes, gestational hypertension, anemia, and preeclampsia (each yes/no), medication use (recorded as generic or trade names, cross-referenced with antidepressant/anxiety medication), current psychiatric disorder (yes/no), gestational age at delivery (weeks and days), and birth weight (kg). Potential covariates were evaluated based on biological plausibility, prior literature, and their associations with psychosocial distress and/or steroid hormone concentrations within the study population.

### Quantification and Statistical Analysis

#### Bivariate analysis

Initially, we compared maternal sociodemographic, prenatal, and steroid characteristics by high-vs. low-distress status to inform covariate selection for modeling. We used χ² tests for categorical variables and Student’s t-tests for continuous variables. We assessed the association of ALLO levels at either pregnancy timepoint with maternal characteristics using ANOVA for categorical predictors and linear trend tests for ordinal predictors. We assessed correlations among biomarkers using Spearman coefficients. These findings, together with known determinants of maternal psychosocial health and steroid biomarkers, informed our modeling approach. We selected covariates a priori based on biological relevance and previous literature rather than solely on statistical significance in bivariate analyses. Because several sociodemographic variables measured related underlying constructs, we selected a parsimonious set of covariates representing distinct domains rather than including all available measures simultaneously.

### Parameterization of the models

For the main analysis, we used linear mixed-effects models which comprise fixed effects, which represent group-level averages (as in traditional regression models) and random effects, which capture individual-level variation about the population average. In these models, binary psychosocial distress status (high vs low) – a fixed effect – is the independent variable of interest and the dependent variable was repeated steroid hormone measures (two values per participant, between 10–34 weeks’ gestation), assessed in separate models. Random effects included random intercepts and slopes. Random intercepts were included in models for all outcomes because it addresses the independence assumption of linear regression, which is otherwise violated by the fact that each participant contributed outcomes across the two study timepoints. In addition to the random intercepts, we considered the need to include random slopes, which allows the change in each steroid outcome across the study timepoints to differ across individuals, using Akaike Information Criterion (AIC). Including random slopes improved model fit for ALLO, ALLO-to-progesterone, ALLO-to-cortisol, and ALLO-to-cortisone ratios and were therefore included in models for these outcomes.

The association of interest from these models is the coefficient for prenatal distress. The β coefficient represents the average difference in the steroid outcome between participants with high versus low prenatal distress across the observed gestational age range (10–34 weeks). In this study, ALLO was specified a priori as the primary outcome/steroid measure of interest, and the other steroid hormones and ratios were assessed as secondary outcomes.

### Multivariable models

Prior to running the multivariable analysis, we assessed effect modification by race/ethnicity using a product term with distress, a standard approach in health equity research.^99–101^ Differential exposure to psychosocial stressors^102^ and structural racism^103^ across groups may influence physiological responses,^104^ and we planned to stratify subsequent analyses if the interaction term was significant. Because we did not observe significant interaction by race/ethnicity, we considered race/ethnicity as a covariate.

For the main analysis, we ran a series of multivariable models that built upon one another, sequentially adjusting for covariates identified a priori and based on bivariate associations. In Model 1, we adjusted for biological and lifestyle factors: maternal age, fetal sex, and prenatal smoking. This model included fetal sex because prior work and our own data showed associations with maternal ALLO concentrations. Model 2 further accounted for sociodemographic characteristics that may be upstream determinants of prenatal distress and pregnancy physiology: use of public assistance programs, nativity, and marital status. Finally, Model 3 included Model 2 covariates plus race/ethnicity. No data were missing for multivariable modeling.

We conducted all analyses in R version 4.5.1 (R Core Team, 2025) using the lme4 and lmerTest packages for mixed-effects modeling and estimates.^105,106^ For all models, we set alpha at 0.05.

