## Supplemental Material for "Associations of Allopregnanolone and Related Steroid Hormones with Prenatal Psychosocial Distress in the Healthy Start Cohort"

**Affiliations:**

### SUPPLEMENTAL MATERIAL:

**Supplemental Table S1.** Bivariate associations of maternal sociodemographic and prenatal characteristics comparing the participants included in this study (n=237) to those Healthy Start participants not included (n=1,173).

| | **Included** | **Not Included** | ***P*-value**[a] |
|---|---|---|---|
| | **n = 237 mean ± SD or N (%)** | **n = 1,173 mean ± SD or N (%)** | |
| **Sociodemographic characteristics** | | | |
| Age (years) | 29 ± 6 | 28 ± 6 | **<0.001** |
| Nulliparous | 91 (38) | 744 (67) | **<0.001** |
| Married/cohabiting | 199 (84) | 931 (80) | 0.20 |
| Use of public assistance program | 72 (31) | 348 (39) | **0.02** |
| Maternal education level | | | **0.01** |
| <High school | 32 (14) | 172 (15) | |
| High school degree or GED | 32 (14) | 227 (19) | |
| Some college or associate’s degree | 45 (19) | 289 (25) | |
| College graduate | 64 (27) | 245 (21) | |
| Graduate degree | 64 (27) | 240 (20) | |
| Annual household income ≥$70,000 | 107 (54) | 353 (38) | **<0.001** |
| Race/ethnicity | | | **<0.001** |
| Non-Hispanic White | 143 (60) | 281 (43) | |
| Hispanic | 54 (23) | 270 (42) | |
| Non-Hispanic Black | 26 (11) | 60 (9.2) | |
| Non-Hispanic Other[b] | 14 (5.9) | 39 (6.0) | |
| Geographic region | | | **0.02** |
| Urban | 219 (96) | 955 (99) | |
| Rural | 8 (3.5) | 11 (1.1) | |
| Born in US | 200 (84) | 1,008 (86) | 0.50 |
| **Prenatal characteristics** | | | |
| Pre-pregnancy BMI, kg/m$^2$ | 25.4 ± 6 | 25.6 ± 6 | 0.30 |
| Fetus is female | 115 (49) | 535 (48) | 0.90 |
| Gestational diabetes mellitus | 11 (5.0) | 44 (4.2) | 0.90 |
| Gestational hypertension | 16 (7.0) | 73 (6.6) | 0.90 |
| Anemia | 44 (19) | 226 (20) | 0.60 |

| | | | |
|---|---|---|---|
| Preeclampsia | 11 (4.8) | 40 (3.6) | 0.40 |
| Smoked during pregnancy | 12 (5.1) | 90 (7.7) | 0.20 |
| Psychiatric disorder | 38 (16) | 204 (18) | 0.50 |
| Use of anti-depressant/anxiety medication during pregnancy | 13 (5.5) | 57 (4.9) | 0.50 |

Data presented as means ± standard deviations or N (%). Bolded values indicate statistical significance at alpha = 0.05.
[a]*P*-values computed by Chi-square or Student T-tests.
[b]Other includes individuals identifying as American Indian or Alaska Native, Hawaiian or Pacific Islander, Asian, or mixed race.

| **Supplemental Table S2.** Effect estimates for high-distress status (β [95% CI] and % Difference [95% CI]) across various sensitivity analyses for allopregnanolone (ALLO) | | | |
|---|---|---|---|
| **Sensitivity Analysis** | **β (95% CI)** | **% Difference (95% CI)** | ***P*-value** |
| ALLO Model 1 (gestational age at blood draw, maternal age, fetal sex, prenatal smoking) | -0.23 (-0.37, -0.09) | -20.57 (-30.87, -8.74) | **<0.001** |
| ALLO Model 1 + Pre-pregnancy BMI (kg/m$^2$) | -0.22 (-0.34, -0.10) | -19.71 (-28.84, -9.41) | **<0.001** |
| ALLO Model 1 + Preeclampsia | -0.24 (-0.39, -0.10) | -21.66 (-31.96, -9.81) | **<0.001** |
| ALLO Model 1 + Anti-depressant/anxiety medication | -0.21 (-0.35, -0.07) | -18.83 (-29.57, -6.44) | **0.004** |

**Supplemental Table S3.** Comparison of high-distress effect estimates (β and %Difference) for all models between the full cohort vs exclusion of the outlier for ALLO, progesterone, and pregnanolone.

| Biomarker or Ratio | Model | Cohort | β (log) [95% CI] | % Difference [95% CI] |
|---|---|---|---|---|
| Allopregnanolone (ALLO) | Unadjusted | Full cohort | -0.23 (-0.36, -0.10) | -20.3 (-29.9, -9.3) |
| | | No-outlier | -0.22 (-0.35, -0.10) | -20 (-29.7, -9.1) |
| | Model 1 | Full cohort | -0.23 (-0.37, -0.09) | -20.6 (-30.9, -8.7) |
| | | No-outlier | -0.23 (-0.37, -0.09) | -20.3 (-30.6, -8.5) |
| | Model 2 | Full cohort | -0.13 (-0.3, 0.01) | -12.4 (-24.4, 1.4) |
| | | No-outlier | -0.13 (-0.28, 0.02) | -12.1 (-24, 1.7) |
| | Model 3 | Full cohort | -0.12 (-0.26, 0.03) | -10.9 (-22.9, 3.1) |
| | | No-outlier | -0.11 (-0.26, 0.03) | -10.5 (-22.5, 3.4) |
| Progesterone | Unadjusted | Full cohort | -0.02 (-0.09, 0.05) | -2 (-8.9, 5.4) |
| | | No-outlier | -0.02 (-0.09, 0.05) | -1.8 (-8.7, 5.5) |
| | Model 1 | Full cohort | 0.01 (-0.07, 0.09) | 1 (-6.6, 9.3) |
| | | No-outlier | 0.01 (-0.07, 0.09) | 1.1 (-6.5, 9.3) |
| | Model 2 | Full cohort | 0.03 (-0.06, 0.11) | 3 (-5.3, 12.1) |
| | | No-outlier | 0.03 (-0.05, 0.11) | 3 (-5.2, 12) |

| | | | | |
|---|---|---|---|---|
| | Model 3 | Full cohort | 0.03 (-0.06, 0.11) | 2.7 (-5.6, 11.8) |
| | | No-outlier | 0.03 (-0.06, 0.11) | 2.8 (-5.5, 11.8) |
| Pregnanolone | Unadjusted | Full cohort | -0.20 (-0.37, -0.03) | -17.9 (-30.6, -2.8) |
| | | No-outlier | -0.19 (-0.36, -0.03) | -17.6 (-30.3, -2.7) |
| | Model 1 | Full cohort | -0.12 (-0.30, 0.06) | -11.4 (-26.1, 6.3) |
| | | No-outlier | -0.12 (-0.30, 0.06) | -11.2 (-25.9, 6.3) |
| | Model 2 | Full cohort | -0.02 (-0.22, 0.17) | -2.2 (-19.4, 18.7) |
| | | No-outlier | -0.02 (-0.21, 0.17) | -2.1 (-19.2, 18.6) |
| | Model 3 | Full cohort | -0.01 (-0.20, 0.19) | -0.7 (-18.2, 20.6) |
| | | No-outlier | -0.01 (-0.20, 0.19) | -0.7 (-18, 20.4) |
| Cortisol | Unadjusted | Full cohort | -0.05 (-0.12, 0.03) | -4.5 (-11.5, 3) |
| | | No-outlier | -0.05 (-0.12, 0.03) | -4.6 (-11.6, 2.9) |
| | Model 1 | Full cohort | -0.05 (-0.14, 0.03) | -5.1 (-12.6, 3) |
| | | No-outlier | -0.05 (-0.14, 0.03) | -5.2 (-12.7, 3) |
| | Model 2 | Full cohort | 0.00 (-0.09, 0.09) | 0.1 (-8.2, 9.1) |
| | | No-outlier | 0.00 (-0.09, 0.09) | 0.1 (-8.2, 9.1) |
| | Model 3 | Full cohort | 0.02 (-0.07, 0.10) | 1.7 (-6.7, 10.7) |

| | | | | |
|---|---|---|---|---|
| | | No-outlier | 0.02 (-0.07, 0.10) | 1.6 (-6.7, 10.7) |
| Cortisone | Unadjusted | Full cohort | 0.04 (-0.02, 0.09) | 3.8 (-1.7, 9.6) |
| | | No-outlier | 0.04 (-0.02, 0.09) | 3.9 (-1.6, 9.6) |
| | Model 1 | Full cohort | 0.03 (-0.03, 0.09) | 3.2 (-2.7, 9.4) |
| | | No-outlier | 0.03 (-0.03, 0.09) | 3.2 (-2.7, 9.4) |
| | Model 2 | Full cohort | 0.04 (-0.03, 0.1) | 3.7 (-2.7, 10.5) |
| | | No-outlier | 0.04 (-0.03, 0.1) | 3.7 (-2.6, 10.5) |
| | Model 3 | Full cohort | 0.04 (-0.02, 0.10) | 4.1 (-2.4, 10.9) |
| | | No-outlier | 0.04 (-0.02, 0.10) | 4.1 (-2.4, 10.9) |
| ALLO-to-Progesterone | Unadjusted | Full cohort | -0.20 (-0.31, -0.09) | -18.1 (-26.4, -9) |
| | | No-outlier | -0.20 (-0.31, -0.09) | -18 (-26.3, -8.8) |
| | Model 1 | Full cohort | -0.23 (-0.35, -0.12) | -20.7 (-29.3, -11.1) |
| | | No-outlier | -0.23 (-0.35, -0.12) | -20.6 (-29.2, -11) |
| | Model 2 | Full cohort | -0.15 (-0.27, -0.03) | -14.1 (-23.9, -3) |
| | | No-outlier | -0.15 (-0.27, -0.03) | -14 (-23.7, -2.9) |
| | Model 3 | Full cohort | -0.13 (-0.25, -0.012) | -12.2 (-22, -1.2) |
| | | No-outlier | -0.13 (-0.25, -0.011) | -12.1 (-21.8, -1.1) |

| | | | | |
|---|---|---|---|---|
| ALLO-to-Pregnanolone | Unadjusted | Full cohort | -0.01 (-0.14, 0.12) | -1.4 (-12.8, 11.4) |
| | | No-outlier | -0.02 (-0.14, 0.12) | -1.5 (-12.8, 11.4) |
| | Model 1 | Full cohort | -0.09 (-0.22, 0.04) | -8.3 (-19.4, 4.3) |
| | | No-outlier | -0.09 (-0.22, 0.04) | -8.3 (-19.4, 4.3) |
| | Model 2 | Full cohort | -0.08 (-0.22, 0.06) | -8 (-19.9, 5.6) |
| | | No-outlier | -0.08 (-0.22, 0.06) | -8 (-19.9, 5.6) |
| | Model 3 | Full cohort | -0.08 (-0.22, 0.06) | -7.7 (-19.7, 6.2) |
| | | No-outlier | -0.08 (-0.22, 0.06) | -7.7 (-19.7, 6.2) |
| ALLO-to-Cortisol | Unadjusted | Full cohort | -0.19 (-0.33, -0.04) | -17 (-28.2, -4.1) |
| | | No-outlier | -0.18 (-0.33, -0.04) | -16.6 (-27.8, -3.8) |
| | Model 1 | Full cohort | -0.19 (-0.35, -0.03) | -17.2 (-29.2, -3.2) |
| | | No-outlier | -0.18 (-0.34, -0.03) | -16.7 (-28.7, -2.7) |
| | Model 2 | Full cohort | -0.15 (-0.32, 0.02) | -13.8 (-27.2, 2.1) |
| | | No-outlier | -0.14 (-0.3, 0.03) | -13.2 (-26.6, 2.6) |
| | Model 3 | Full cohort | -0.15 (-0.32, 0.02) | -13.5 (-27, 2.4) |
| | | No-outlier | -0.14 (-0.31, 0.03) | -12.9 (-26.4, 3.1) |
| ALLO-to-Cortisone | Unadjusted | Full cohort | -0.26 (-0.39, -0.13) | -22.6 (-32.1, -11.8) |

| | | | | |
|---|---|---|---|---|
| | | No-outlier | -0.25 (-0.38, -0.12) | -22.4 (-31.9, -11.6) |
| | Model 1 | Full cohort | -0.25 (-0.40, -0.11) | -22.4 (-32.7, -10.6) |
| | | No-outlier | -0.25 (-0.39, -0.11) | -22.1 (-32.4, -10.3) |
| | Model 2 | Full cohort | -0.16 (-0.31, -0.01) | -14.7 (-26.6, -0.8) |
| | | No-outlier | -0.16 (-0.30, -0.01) | -14.3 (-26.2, -0.5) |
| | Model 3 | Full cohort | -0.14 (-0.29, 0.01) | -13.2 (-25.2, 0.7) |
| | | No-outlier | -0.14 (-0.29, 0.01) | -12.8 (-24.8, 1) |
| Cortisol/Cortisone | Unadjusted | Full cohort | -0.08 (-0.14, -0.03) | -8 (-13.3, -2.5) |
| | | No-outlier | -0.09 (-0.14, -0.03) | -8.2 (-13.4, -2.7) |
| | Model 1 | Full cohort | -0.08 (-0.15, -0.02) | -8 (-13.7, -2) |
| | | No-outlier | -0.09 (-0.15, -0.02) | -8.1 (-13.8, -2.1) |
| | Model 2 | Full cohort | -0.04 (-0.10, 0.03) | -3.5 (-9.8, 3.2) |
| | | No-outlier | -0.04 (-0.10, 0.03) | -3.6 (-9.8, 3.1) |
| | Model 3 | Full cohort | -0.02 (-0.09, 0.04) | -2.3 (-8.6, 4.4) |
| | | No-outlier | -0.02 (-0.09, 0.04) | -2.4 (-8.6, 4.3) |

**Abbreviations:** CI – confidence interval.

**Model 1:** Adjusts for gestational age at blood draw for biomarker measurement, maternal age at enrollment, fetal sex, and prenatal smoking.
**Model 2:** Model 1 + use of public assistance programs, nativity (U.S.-born yes/no), and marital status.
**Model 3:** Model 2 + race/ethnicity.

Biomarker concentrations were modeled on the log natural scale. % difference = (exp(estimate) – 1) *100.

**Supplemental Table S4.** Intra- and Inter-day accuracy and precision from a three-day abbreviated validation of the steroid hormone assay on the analytical platform used in the present study. Quality control samples ($n \geq 5$/day) were analyzed across three independent days at low, medium, and high concentrations. Accuracy is reported as percent nominal concentration recovered, and precision is reported as coefficient of variation (CV%). The abbreviated validation was performed to verify assay performance following transfer of the previously validated method to the current analytical platform.

| | Intra-day ranges | | Inter-day ranges | |
|---|---|---|---|---|
| | Accuracy | Precision | Accuracy | Precision |
| | % | %CV | % | %CV |
| ALLO | 86.4 - 115.5 | 0.91 - 14.46 | 97.2 - 108.1 | 7.66 - 12.72 |
| Pregnanolone | 90.5 - 115.3 | 0.99 - 14.6 | 97.1 - 110.4 | 6.81 - 8.87 |
| Progesterone | 101.9 - 118.5 | 1.14 - 7.23 | 110.3 - 112.7 | 4.41 - 7.16 |
| Cortisone | 88 - 115.6 | 0.86 - 8.11 | 86.6 - 111.1 | 4.01 - 6.86 |
| Cortisol | 93.2 - 116.2 | 1.56 - 7.81 | 106.6 - 114.9 | 5.28 - 8.16 |

**Supplemental Table S5.** Multiple reaction monitoring (MRM) transitions and optimized mass spectrometer acquisition parameters used for quantification of steroid hormones and internal standards on the AB Sciex API6500 tandem mass spectrometer.

| Compound | Q1 | Q3 | DP (V) | CE (V) | CXP (V) |
|---|---|---|---|---|---|
| Allopregnanolone | 301.2 | 283.5 | 50 | 19 | 12 |
| Pregnanolone | 301.2 | 189.2 | 50 | 29 | 12 |
| Progesterone | 315.3 | 109 | 88 | 33 | 7 |
| Cortisone | 361 | 163.3 | 90 | 33 | 12 |
| Cortisol | 363.1 | 121 | 76 | 30 | 8 |
| Allopregnanolone-d5 | 306.2 | 288.2 | 50 | 19 | 12 |
| Cortisone-d8 | 369.2 | 168.4 | 102 | 31 | 14 |
| Progesterone-d9 | 324.4 | 100.1 | 116 | 45 | 14 |
| Cortisone-d4 | 367.4 | 121 | 110 | 36 | 22 |

Q1 and Q3 denote precursor and product ion mass-to-charge ratios (m/z); CE, collision energy; CXP, collision cell exit potential; DP, delustering potential.

**Supplemental Figure S1.** Heat maps of biomarker correlations using Spearman (Rho) at (**A**) first blood sample (median 17 weeks' gestation) and (**B**) second blood sample (median 27 weeks' gestation) among 237 pregnant individuals in the Healthy Start cohort.

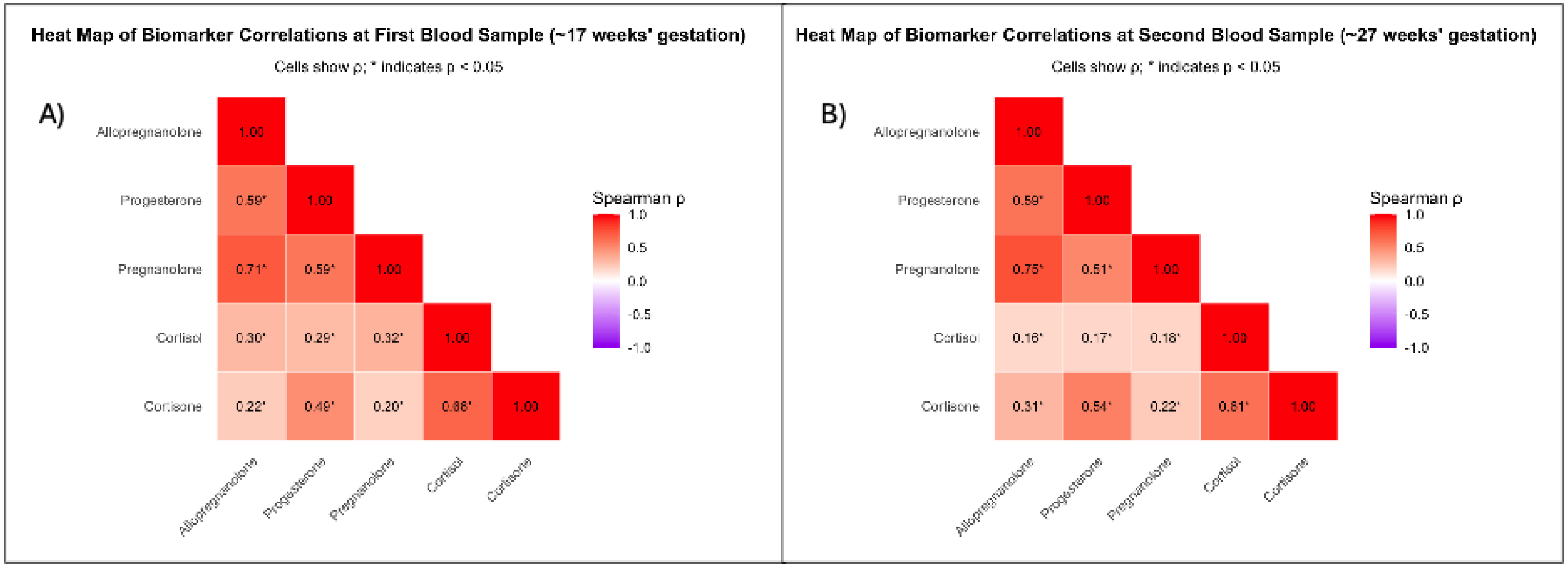

**Supplemental Figure S2.** Representative extracted ion chromatograms for the quantification of allopregnanolone (ALLO, 3α-hydroxy-5α-pregnan-20-one), progesterone, pregnanolone (3α- hydroxy-5β-pregnan-20-one), cortisol, and cortisone using a liquid chromatography–tandem mass spectrometry (LC–MS/MS) assay validated according to the FDA/ICH M10 guideline.[1] The quantified concentration corresponding to each representative chromatogram is as follows: pregnanolone = 4.3 ng/mL, ALLO = 8.9 ng/mL, progesterone = 39.1 ng/mL, cortisone = 28.1 ng/mL, and cortisol = 160.1 ng/mL.

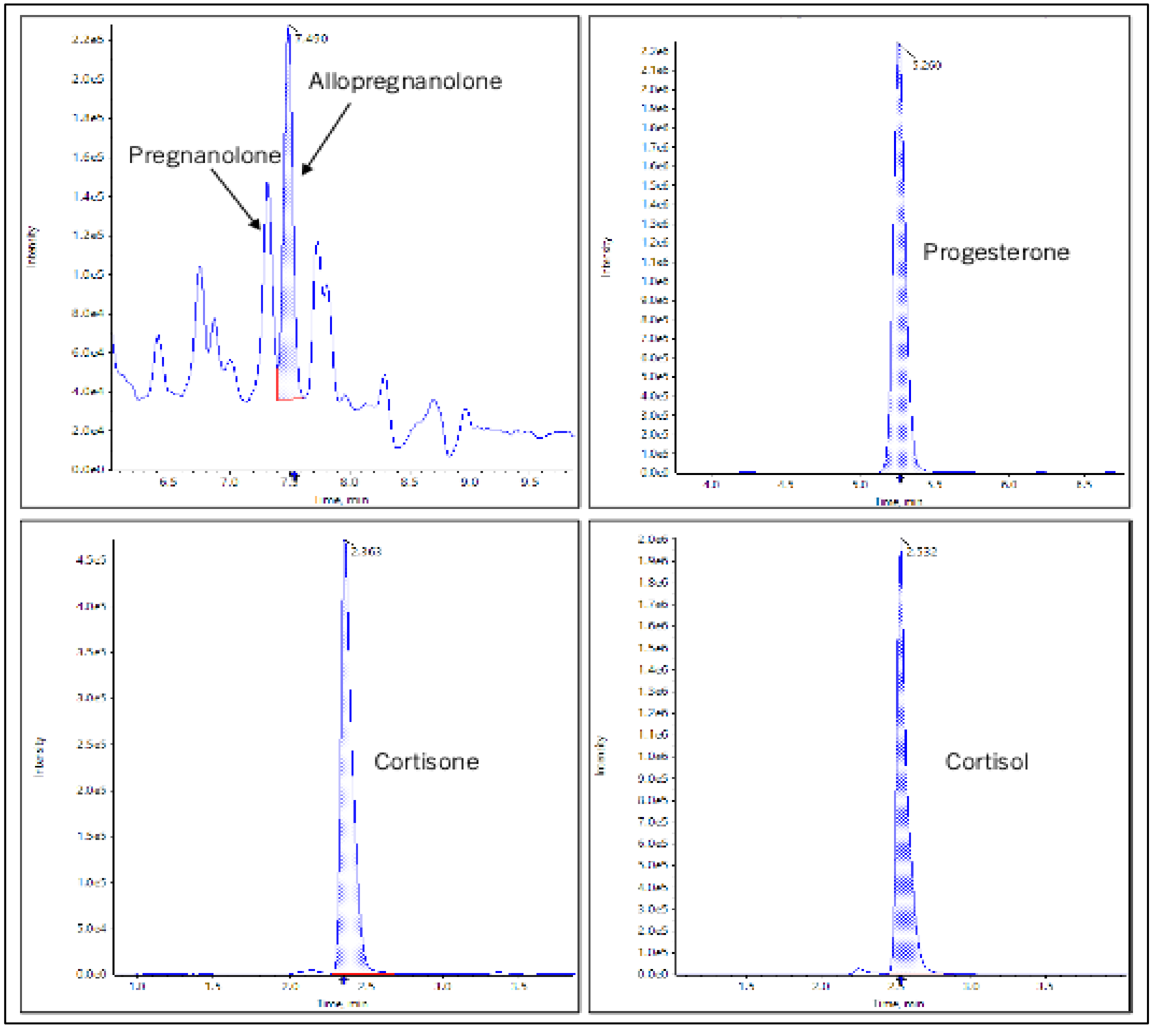

**Figure S3.** Connections and positions of the switching valve for online extraction (on the left) and backflush (on the right).

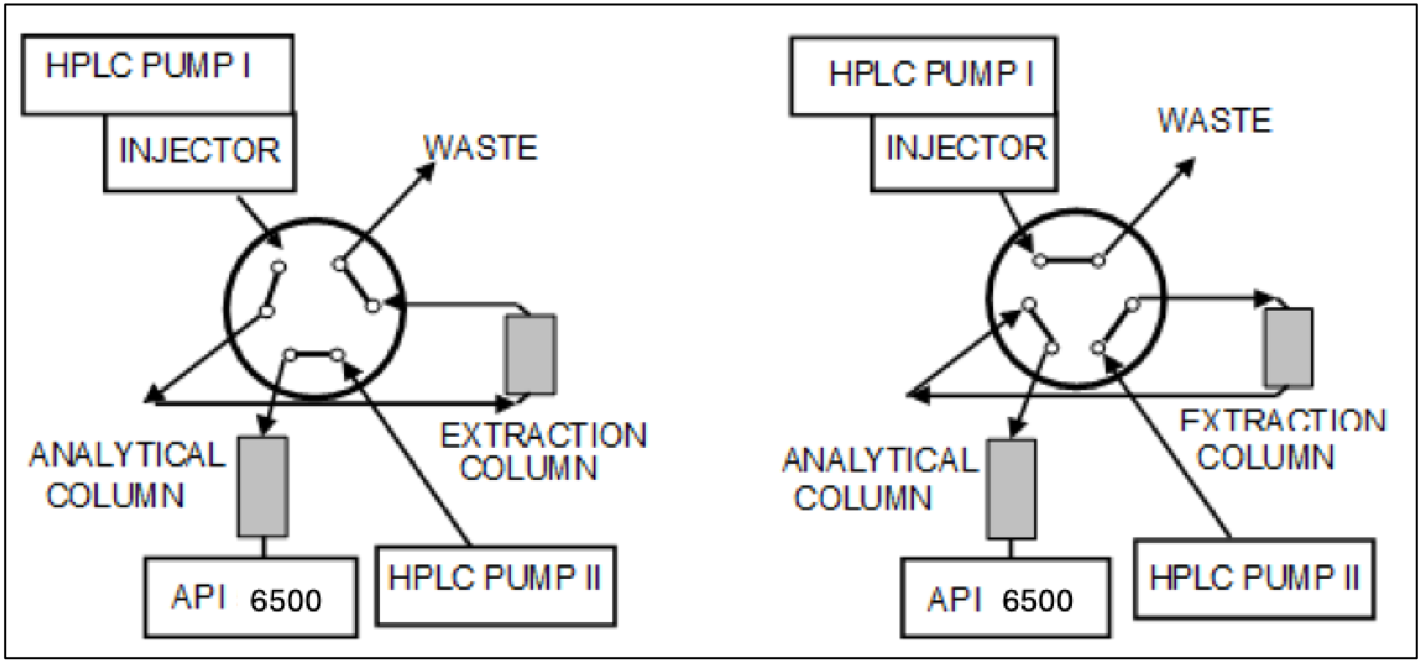

**Supplemental Methods:**

HPLC-MS/MS parameters were the same as the original published assay.[1] Briefly, frozen serum samples were thawed on ice and 100 µL was aliquoted into microcentrifuge tubes. 400 µL of pure methanol containing 25 ng/mL deuterated internal standards (allopregnanolone-d5, progesterone-d9, cortisol-d4, cortisone-d8, and pregnanolone-d5) was added to precipitate proteins. Allopregnanolone-d5, progesterone-d9, cortisol-d4, and cortisone-d8 were purchased from Sigma-Aldrich (MilliporeSigma), whereas pregnanolone-d5 was purchased from Toronto Research Chemicals (Ontario, Canada). Analytical standards for allopregnanolone and cortisone were obtained from Toronto Research Chemicals, cortisol and progesterone from Cayman Chemical, and pregnanolone from Sigma-Aldrich. Samples were vortexed for 5 minutes and centrifuged at 25,000×g and 4 °C for 10 minutes. The supernatant was transferred into HPLC vials and loaded into the autosampler. Chromatographic separation was performed on an Agilent HPLC system (two binary pumps, thermostated column compartment with an integrated six-port switching valve, and Leap HTC-xt autosampler). The HPLC system was coupled to an AB Sciex API6500 tandem mass spectrometer (Sciex, Concord, ON, Canada) with a turbo ion spray source operated in positive ionization mode. Online extraction was performed using a previously developed column-switching and backflush methodology.[2] This approach concentrates analytes while removing matrix components prior to analytical separation, resulting in improved chromatographic performance and assay throughput.

For online extraction, one hundred microliters (100 µL) of the extracted samples, calibrators and QC samples were injected and loaded onto an Agilent Zorbax XDB-C18, 50 x 2.1 mm (5 µm particle size) HPLC extraction column at 85% LC-MS grade water with 0.1% formic acid and 15% LC-MS grade methanol with 0.1% formic acid at a flow rate increasing from 0.8 mL/minute to 1.5 mL/min within one minute. The retained analytes were then backflushed using a six-port switching valve onto the analytical column (Agilent Poroschell 120, 50 x 4.6 mm, 2.7 µm particle size, EC-C18 material, Agilent Technologies, Santa Clara, CA, USA). The connections and positions of the 6-port switching valve (Rheodyne, Cotati, CA, USA) are shown in **Supplemental Figure S3**.

The API6500 tandem mass spectrometer was used in the positive electrospray ionization mode (+ESI) at an ionization voltage of 5500V in combination with multiple reaction monitoring (for more details, please see **Supplemental Table S5**).

Before analyzing study samples, we conducted a three-day abbreviated validation on this platform pursuant to clinical and industry guidelines.[3] The purpose of this validation was to confirm assay performance following transfer of the previously validated method to a new analytical platform. Acceptance criteria for intra- and inter-day imprecision (coefficient of variation) were ≤20% for all calibrators (≤25% at the lower limit of quantification [LLOQ]) and trueness had to be within ±20% of nominal values (±25% at the LLOQ). Acceptance criteria for calibration curve fit were $r^2 \geq 0.99$ and at least 75% of calibrators had to meet trueness criteria. Quality control (QC) samples in low, medium, and high concentration ranges for each analyte were included. Accuracy and precision of QC samples were within ± 15% of nominal concentrations (± 20% at the LLOQ). Matrix effects and recovery were not re-evaluated during the abbreviated platform validation because the extraction procedure and sample preparation were unchanged from the original validated assay method. On the current platform, the working ranges were 0.39–100 ng/mL for allopregnanolone and pregnanolone, 1.56–400 ng/mL for progesterone and cortisone, and 3.91–1000 ng/mL for cortisol. In our original assay validation, all analytes remained within acceptance criteria after three freeze–thaw cycles and after three months of storage at −80 °C. Long-term frozen-storage stability has been demonstrated for steroid hormones, including progesterone and cortisol, and similar stability is expected for the structurally related neuroactive steroids measured here.[4,5]
